# Vitamin D Supplements for Prevention of COVID-19 or other Acute Respiratory Infections: a Phase 3 Randomised Controlled Trial (CORONAVIT)

**DOI:** 10.1101/2022.03.22.22271707

**Authors:** David A. Jolliffe, Hayley Holt, Matthew Greenig, Mohammad Talaei, Natalia Perdek, Paul Pfeffer, Giulia Vivaldi, Sheena Maltby, Jane Symons, Nicola L. Barlow, Alexa Normandale, Rajvinder Garcha, Alex G. Richter, Sian E. Faustini, Christopher Orton, David Ford, Ronan A. Lyons, Gwyneth A. Davies, Frank Kee, Christopher J. Griffiths, John Norrie, Aziz Sheikh, Seif O. Shaheen, Clare Relton, Adrian R. Martineau

## Abstract

**OBJECTIVES:** To determine whether population-level implementation of a test-and-treat approach to correction of sub-optimal vitamin D status (25-hydroxyvitamin D [25(OH)D] <75 nmol/L) influences risk of all-cause acute respiratory infection (ARI) or coronavirus disease 2019 (COVID-19).

**DESIGN:** Phase 3 open-label randomised controlled trial (CORONAVIT) utilising trials-within-cohorts (TwiCs) methodology.

**SETTING:** United Kingdom.

**PARTICIPANTS:** 6200 adults aged 16 years or older, who were not already taking vitamin D supplements at baseline.

**INTERVENTIONS:** Offer of a postal finger-prick test of blood 25(OH)D concentration with provision of a 6-month supply of higher-dose vitamin D (3200 IU/day, n=1550) or lower-dose vitamin D (800 IU/day, n=1550) to those with blood 25(OH)D concentration <75 nmol/L, vs. no offer of testing or supplementation (n=3100). Follow-up was from 17^th^ December 2020 to 16^th^ June 2021.

**MAIN OUTCOME MEASURES:** The primary outcome was the proportion of participants experiencing at least one doctor- or swab test-confirmed ARI of any cause. Secondary outcomes included the proportion of participants developing swab test-confirmed COVID-19. Logistic regression was used to calculate odds ratios and associated 95% confidence intervals.

**RESULTS:** Of 3100 participants offered 25(OH)D testing, 2958 (95.4%) accepted, and 2690 (86.8%) had 25(OH)D <75 nmol/L and were sent vitamin D supplements (1356 higher-dose, 1334 lower-dose). 76 (5.0%) vs. 87 (5.7%) vs. 136 (4.6%) participants in higher-dose vs. lower-dose vs. no offer groups experienced at least one ARI of any cause (odds ratio [OR] for higher-dose vs. no offer 1.09, 95% CI 0.82-1.46; lower-dose vs. no offer 1.26, 0.96-1.66). 45 (3.0%) vs. 55 (3.6%) vs. 78 (2.6%) participants in higher-dose vs. lower-dose vs. no offer groups developed COVID-19 (OR for higher-dose vs. no offer 1.13, 0.78-1.63; lower-dose vs. no offer 1.39, 0.98-1.97).

**CONCLUSIONS:** Among adults with a high baseline prevalence of sub-optimal vitamin D status, implementation of a population-level test-and-treat approach to vitamin D replacement did not reduce risk of all-cause ARI or COVID-19.

**TRIAL REGISTRATION:** ClinicalTrials.gov no. NCT04579640

**SUMMARY BOX:** *What is already known on this topic?:* Vitamin D metabolites support innate immune responses to severe acute respiratory syndrome coronavirus 2 (SARS-CoV-2) and other respiratory pathogens. Sub-optimal vitamin D status (25-hydroxyvitamin D <75 nmol/L) associates with increased susceptibility to all-cause acute respiratory infections (ARI) and coronavirus disease 2019 (COVID-19). Phase 3 randomised controlled trials of vitamin D to prevent COVID-19 have not yet reported.

*What this study adds:* This phase 3 randomised controlled trial, including 6200 participants, shows that implementation of a population-level test-and-treat approach to oral vitamin D replacement at a dose of 800 IU or 3200 IU per day did not reduce risk of all-cause ARI or COVID-19 among adults with a high baseline prevalence of sub-optimal vitamin D status.

## INTRODUCTION

The coronavirus disease 2019 (COVID-19) pandemic has refocused attention on strategies to prevent acute respiratory infection (ARI). Although vaccination against severe acute respiratory syndrome coronavirus 2 (SARS-CoV-2) represents the mainstay for disease control, its effectiveness at a global level is compromised by factors including cost, availability, vaccine hesitancy, vaccine failure and vaccine escape.^1–3^ Complementary, low-cost approaches to enhance immunity to SARS-CoV-2 and other pathogens causing ARI are needed.

Vitamin D metabolites have long been recognised to support diverse innate immune responses to respiratory viruses and bacteria, while simultaneously regulating immunopathological inflammation.^4–6^ The vitamin D-inducible antimicrobial peptides cathelicidin LL-37 and human beta defensin 2 have both been shown to bind SARS-CoV-2 spike protein and inhibit binding to Angiotensin Converting Enzyme 2, its cellular receptor.^7^ ^8^ Longitudinal studies investigating potential associations between higher vitamin D status or vitamin D supplement use and reduced risk of COVID-19 or SARS-CoV-2 infection have yielded mixed results, with some reporting protective associations and others reporting null or negative associations;^9–14^ meta-analyses including these and other observational studies report protective associations overall.^15^ ^16^ Findings of randomised controlled trials (RCTs) of vitamin D supplementation to prevent ARIs caused by pathogens other than SARS-CoV-2 have also been heterogeneous.^17–21^ Meta-analysis of these and other RCTs shows a small, but statistically significant, protective effect that is strongest where modest daily doses of vitamin D (400-1000 IU) are given for periods of up to one year.^22^ Very recently, a Phase 2 RCT conducted in 321 Mexican healthcare workers reported a large protective effect of daily oral vitamin D supplementation against COVID-19 (relative risk 0.23, 95% CI 0.09 to 0.55).^23^ However, Phase 3 clinical trials of prophylactic vitamin D to reduce incidence and severity of COVID-19 are lacking, as are studies comparing effectiveness of different doses of vitamin D supplementation for the prevention of ARIs of any cause among adults. There is also a lack of studies designed to evaluate the effectiveness of practical approaches to identification and treatment of vitamin D deficiency at scale in the general population to improve health outcomes. We therefore established a Phase 3 pragmatic RCT (CORONAVIT) to evaluate the effectiveness of a ‘test-and-treat’ approach to identification and treatment of vitamin D insufficiency for prevention of COVID-19 and other ARIs in U.K. adults from December 2020 to June 2021 - a period when COVID-19 incidence was high and COVID-19 vaccine coverage was initially low.

## METHODS

### STUDY DESIGN

We conducted a three-arm parallel open-label individually-randomised controlled trial nested within the population-based COVIDENCE UK cohort study, using ‘trial-within-cohort’ methodology.^24^ COVIDENCE UK was established to determine risk factors for incident COVID-19;^12^ ^13^ to characterise the natural history of COVID-19; to evaluate impacts of COVID-19 on physical and mental health and economic vulnerability;^25^ and to provide a platform from which to conduct clinical trials of interventions to reduce incidence and/or severity of ARI. The study was launched on 1st May 2020 and closed to recruitment on 6th October 2021; a total of 19,981 participants enrolled. The CORONAVIT trial – the first RCT to be nested within the cohort - was sponsored by Queen Mary University of London, approved by the Queens Square Research Ethics Committee, London, UK (ref 20/HRA/5095) and registered with ClinicalTrials.gov (NCT04579640) on 8^th^ October 2020, i.e. before enrolment of the first participant on 28^th^ October 2020.

### PARTICIPANTS

Trial participants were drawn from participants in the COVIDENCE UK longitudinal study, who provided information for the following variables using an on-line questionnaire that was completed at enrolment: age, sex, ethnic origin, postal address, highest educational level attained, occupational status, weight, height, past history of SARS-CoV-2 infection or COVID-19, COVID-19 vaccination status, longstanding health conditions, tobacco smoking history, alcohol consumption, intake of micronutrient supplements, hours spent outdoors, and dietary consumption of oily fish and red meat. Inclusion criteria were current residence in the UK, age 16 years or more at screening, enrolment in the COVIDENCE UK longitudinal cohort study (NCT04330599), and on-line provision of informed consent. Exclusion criteria were taking vitamin D supplements, digoxin, alfacalcidol, calcitriol, dihydrotachysterol or paricalcitol; known diagnosis of sarcoidosis, primary hyperparathyroidism, nephrolithiasis, or renal failure requiring dialysis; known allergy to any ingredient in the study capsules; and known pregnancy.

### RANDOMISATION

A randomly-selected subset of 6200 cohort participants who were assessed as eligible on the basis of data from their enrolment questionnaire, and who reported no supplemental vitamin D intake at baseline, were individually randomised by the trial statistician using a computer program (Stata v14.2) to receive an offer of a postal vitamin D test with supply of 3200 IU vitamin D/day if their blood 25(OH)D concentration was found to be less than 75 nmol/L (‘higher-dose offer group’) vs. the same testing offer with supply of 800 IU vitamin D/day if 25(OH)D was less than 75 nmol/L (‘lower-dose offer group’) vs. no offer of vitamin D testing or supplementation (‘no offer group’), with a 1:1:2 allocation ratio. To implement the randomisation, the study statistician downloaded unique identifiers of all participants in the COVIDENCE UK longitudinal study whose questionnaire responses indicated that they were eligible to participate in the CORONAVIT trial. Stata software was then used to randomly select 6200 of those reporting the lowest supplemental vitamin D intake, and to randomly assign one of the labels ‘higher-dose offer’, ‘lower-dose offer’ or ‘no offer’ to each of them in a 1:1:2 ratio in blocks of ten, such that 1550 participants were randomised to each of the offers and 3100 participants were randomised to no offer. Treatment allocation was not concealed, and randomisation was not stratified.

### INTERVENTION

Consenting participants randomised to either intervention arm of the trial were posted a blood spot testing kit for determination of 25(OH)D concentrations in capillary blood, as previously described.^26^ Those found to have a 25(OH)D concentration of less than 75 nmol/L were then posted a 6-month supply of capsules containing either 800 IU or 3200 IU vitamin D_3_, according to their allocation. The 75 nmol/L threshold was utilised on the grounds that this is widely considered to discriminate between those with optimal vs. sub-optimal vitamin D status.^27–29^ We adopted a ‘test-and-treat’ approach in order to avoid the potential risk of inducing hypervitaminosis D by providing higher-dose supplementation to individuals who had baseline 25(OH)D concentrations >75 nmol/L, and in response to feedback from our patient and public involvement group who advised that participants might be better motivated to adhere to supplementation if they knew that their baseline 25(OH)D concentration was sub-optimal.

Participants were supplied with D-Pearls capsules of either strength, manufactured by Pharma Nord Ltd (Vejle, Denmark), unless they expressed a preference for a vegetarian or vegan supplement, in which case they were supplied with Pro D_3_ vegan capsules manufactured by Synergy Biologics Ltd (Walsall, UK). Participants with 25(OH)D concentrations of 75 nmol/L or more at initial testing were offered a second postal vitamin D test 2 months after the first test: those whose second 25(OH)D concentration was found to be less than 75 nmol/L were offered a postal supply of supplements as above. Participants receiving study supplements were instructed to take one capsule per day until their supply was exhausted. Administration of study supplements was not supervised.

### FOLLOW-UP ASSESSMENTS

Follow-up was from 17^th^ December 2020 to 16^th^ June 2021. All participants were emailed links to follow-up on-line questionnaires at monthly intervals that captured the following information: incident test-positive or doctor-diagnosed acute respiratory infections (ARIs) including COVID-19, hospitalisation for treatment of ARIs including COVID-19, requirement for ventilatory support, prescription of antibiotic treatment for ARI, incidence of acute exacerbations of asthma or COPD and other adverse events, uptake of COVID-19 vaccination, presence or absence of long-term COVID- 19 symptoms at 4-weeks post-diagnosis and at 6-month follow-up (participants with test-positive COVID-19), MRC dyspnoea score,^30^ FACIT Fatigue Score^31^ and Post-Covid Physical Health Symptom Score ^32^ at 6-month follow-up (participants with test-positive COVID-19). This score was based on responses to questions that participants were asked about the following 16 symptoms they might be experiencing post-Covid: excessive shortness of breath, cough, unusual tiredness or fatigue, problems with sleeping, memory problems, difficulty concentrating, joint or muscle pain, problems with taste or smell, diarrhoea, stomach (abdominal) pains, changes to voice, hair loss, unusual racing of the heart, light-headedness or dizziness, unusual sweating, and ringing or buzzing in the head or in one or both ears that lasts for more than five minutes at a time. For each symptom, participants were given the following response options: ‘No’ (1 point), ‘Yes but improving’ (2 points), ‘Yes but not improving or worsening’ (3 points), or ‘Yes and worsening’ (4 points), with the sum being the Symptom Score (lower scores signifying fewer post-Covid symptoms).

Details of incident swab test- or doctor-confirmed ARIs, hospitalisations and deaths were also captured via electronic linkage to routinely collected medical record data. End-trial postal vitamin D testing was offered to a randomly selected subset of 1600 participants who received study supplements (800 vs. 800 randomised to higher- vs. lower-dose offers, respectively) and 400 adults who were randomised to no offer. Participants randomised to no offer who were found to have end-trial 25(OH)D concentrations <50 nmol/L were posted a 60-day supply of capsules each containing 2500 IU vitamin D_3_ (Cytoplan Ltd).

Every monthly questionnaire contained the following advice to encourage participants with COVID-19 symptoms to engage with testing services: “If you currently have symptoms of coronavirus (a high temperature, a new, continuous cough or loss of or altered sense of smell or taste), call NHS111 or visit https://www.nhs.uk/conditions/coronavirus-covid-19/ for more information.” This wording was identical for questionnaires sent to participants randomised to intervention vs. no-offer groups. In addition to monthly questionnaires detailed above, a link to an online adherence questionnaire was sent to all participants randomised to either offer on 31^st^ March 2021. This captured information regarding frequency of study supplement use.

### LABORATORY TESTING

25(OH)D assays were performed by Black Country Pathology Services, located at Sandwell General Hospital, West Bromwich, UK; this laboratory participates in the UK NEQAS for Vitamin D and the Vitamin D External Quality Assessment Scheme (DEQAS) for serum 25(OH)D. Concentrations of 25(OH)D_3_ and 25(OH)D_2_ were determined in dried blood spot eluates using liquid chromatography tandem mass spectrometry (Acquity UPLC-TQS or TQS-Micro Mass Spectrometers, Waters Corp., Milford, MA) after derivatisation and liquid–liquid extraction as previously described^26^ and summed to give total 25(OH)D concentrations. Very good overall agreement between blood spot and plasma 25(OH)D concentrations in paired capillary and venous samples using this blood spot method has been observed,^26^ demonstrating a minimal overall bias of -0.2% with a bias range of -16.9 to 26.7%. The following reference ranges for total 25(OH)D concentrations were used for the study: deficient or sub-optimal <75.0 nmol/L; adequate 75.0-220.0 nmol/L; high >220.0 nmol/L. The between-day coefficients of variation were: 11.1% at 16.9 nmol/L, 8.2% at 45.5 nmol/L, 6.9% at 131.7 nmol/L and 7.0% at 222.2 nmol/L for 25(OH)D_3_ and 13.7% at 18.1 nmol/L, 7.5% at 42.7 nmol/L and 6.4% at 127.3 nmol/L for 25(OH)D_2_. The mean bias of dried blood spot vs. serum 25(OH)D_3_ concentrations over the period 2018 to 2021 was 4.0% and the limits of quantitation (LOQ) were 7.5 nmol/L for 25(OH)D_3_ and 2.8 nmol/L for 25(OH)D_2_.

### OUTCOMES

The primary outcome was the proportion of participants developing at least one swab test- or doctor-confirmed ARI of any cause. These events were captured either via self-report using structured monthly on-line questionnaires and/or via linkage to one or more of the following databases containing routinely collected virology test results and medical record data from primary, secondary and tertiary healthcare facilities: the UK Office for National Statistics (ONS) mortality database, the UK National Health Service Hospital Episode Statistics (HES) database, the COVID-19 Hospitalisation in England Surveillance System (CHESS) database, the General Practice Extraction Service (GPES) database and the Second Generation Surveillance Service (SGSS) database, a national laboratory reporting system used to capture routine laboratory data on infectious diseases. The wording of questions from monthly follow-up questionnaires that were used to capture self-reported swab test- or doctor-confirmed ARI, along with the algorithms used to define these events, are displayed in Table S1, Supplementary Appendix. Episodes of swab test- or doctor-confirmed ARI documented in medical records were captured via computerised searches based on International Classification of Diseases, 10^th^ revision (ICD-10) codes for ARI, listed in Table S2, Supplementary Appendix.

Secondary efficacy outcomes were the proportion of all participants developing COVID-19 confirmed by reverse transcription polymerase chain reaction (RT-PCR) or antigen testing; the proportion of all participants hospitalised for treatment of COVID-19; the proportion of participants hospitalised for treatment of COVID-19 who required ventilatory support; the proportion of all participants dying of COVID-19; the proportion of participants developing test-confirmed COVID-19 who reported symptoms lasting more than four weeks; the proportion of participants developing test-confirmed COVID-19 who reported on-going symptoms at the end of the study; mean values for the MRC dyspnoea score^30^, the FACIT Fatigue Scale score^31^ and the Post-Covid Physical Health Symptom Score^32^ among participants developing test-confirmed COVID-19 who reported on-going symptoms at the end of the study; the proportion of all participants prescribed one or more courses of antibiotics for treatment of ARI of any cause; the proportion of all participants hospitalised for treatment of ARI of any cause; the proportion of all participants dying of ARI of any cause; the proportion of participants with asthma or COPD developing at least one severe acute exacerbation; and mean end-study 25(OH)D concentrations in the subset of participants for whom this was measured. Safety outcomes were incidence of death, serious adverse events, adverse events leading to discontinuation of study medication, and other monitored safety conditions: hypercalcemia (serum corrected calcium concentration >2.65 mmol/L), hypervitaminosis D (25[OH]D concentration >220 nmol/L) and nephrolithiasis.

### SAMPLE SIZE CALCULATION

We used https://mjgrayling.shinyapps.io/multiarm/^33^ to calculate that a total of 6200 participants would need to be randomised to detect a 20% reduction in the proportion of participants meeting the primary outcome with 84% marginal power^34^ and 5% type 1 error rate, assuming a 20% risk of experiencing at least one swab test- or doctor-confirmed ARI in the no offer group at six months, 25% loss to follow-up, and a 1:1:2 ratio of participants randomised to higher-dose, lower-dose or no offer, respectively.

## STATISTICAL METHODS

Statistical analyses were performed using Stata 14.2 (College Station, TX). Primary analyses were performed according to intention-to-treat (ITT), with pair-wise comparisons made between each intervention arm and the no offer arm. All randomised participants who had data available for at least one follow-up time point were included in the ITT analysis. Proportions of participants experiencing dichotomous study endpoints were compared between intervention vs. no offer arms using logistic regression, with treatment effects presented as odds ratios (ORs) with 95% confidence intervals (CIs). Adjustment for multiple testing for the primary outcome (arising from the fact that two comparisons were being made) used Dunnett’s test^35^ with a critical P-value threshold of 0.027. Continuous outcomes were compared between intervention vs. no offer arms using unpaired Student’s t-tests, with treatment effects presented as mean differences with 95% confidence intervals.

We pre-specified sub-group analyses comparing the effect of the intervention on major outcomes in individuals who received at least one dose of COVID-19 vaccine during follow-up vs. those who did not, and in those with lower vs. higher actual or predicted baseline 25(OH)D concentrations. For each of these sub-group analyses, we pre-specified testing for interactions between allocation and the potential effect-modifier of interest. Additionally we conducted a post hoc exploratory Cox regression analysis to determine effects of allocation before vs. after administration of a first dose of COVID-19 vaccine. Predicted baseline 25(OH)D concentrations for participants in the no offer arm were calculated using published methodology^36^ using baseline 25(OH)D values of participants randomised to either intervention arm using a random forest regression model ^37^ with 11 features (age, sex, body mass index [BMI], ethnicity, portions of read meat per week, portions of oily fish per week, use of multivitamin supplements [yes/no], use of vitamin D supplements [yes/no], use of cod liver oil supplements [yes/no], sunshine hours per week, and latitude). Sunshine hours per week were obtained from HadUK’s 2019 5km gridded climate measurements using the mean value of sunshine hours per week in the period October-December 2019 in the postcode grid square for each participant. For age, BMI, sunshine hours per week, and latitude, participants with missing data were imputed with the median value of that covariate among participants with non-missing data. For all other covariates, participants with missing data were imputed with the mode of that covariate among participants with non-missing data. Using a training set comprising 75% of all instances, we conducted a grid search over hyperparameter values using a 5-fold cross-validation procedure. The following hyperparameter values were searched: split criterion (mean squared error and mean absolute error), maximum tree depth (3, 4, 5, 6), bootstrap sample size (0.5, 0.65, 0.8) and number of trees (250, 500, 1000). Hyperparameter combinations were evaluated using the mean gamma deviance over all 5 folds of the cross-validation split. The optimal hyperparameter combination produced a mean gamma deviance of 0.151 across all validation folds. To verify the generalisation accuracy of the model trained with these hyperparameters, we evaluated the model on a test set comprising 25% of all instances, which yielded a gamma deviance of 0.141, showing that the out-of-sample performance observed in the cross-validation was indicative of test set performance. The final model was trained on all instances with available data and used to impute baseline 25(OH)D concentrations for participants randomised to the no offer arm of the trial. Imputed baseline 25(OH)D values for participants randomised to the no offer arm of the trial were then compared to measured baseline 25(OH)D values for participants randomised to either intervention arm (Figure S1, Supplementary Appendix): the distribution of imputed 25(OH)D concentrations in the ‘no offer’ arm at baseline (range 61.2 nmol/L, s.d. 8.2 nmol/L) was constrained compared to that of measured 25(OH)D concentrations in the lower-offer arm (range 169.3 nmol/L, s.d. 17.8 nmol/L) and the higher-offer arm (range 111.7 nmol/L, s.d. 16.2). This finding called the validity of the imputation into question, which precluded conduct of sub-group analyses by baseline 25(OH)D concentration.

A sensitivity analysis was also pre-specified, which excluded data from participants in the intervention arms who reported that they took vitamin D capsules ‘less than half the time’ as well as those in the no offer arm who reported any intake of supplemental vitamin D during follow-up. Two post hoc exploratory analyses were also performed in response to reviewer comments. The first compared mean end-study 25(OH)D concentrations of control arm participants who contributed data to the ITT analysis but not to the sensitivity analysis (i.e. participants randomised to no offer who reported use of off-trial vitamin D supplements) vs. those who contributed data to both ITT & sensitivity analyses (i.e. participants randomised to no offer who reported no use of off-trial vitamin D supplements). The second evaluated the effect of allocation in the subset of participants who completed all six follow-up questionnaires.

No interim analysis was planned or performed due to the short duration of the trial, and the fact that linkage to routinely collected health data was not done until the end of the trial. However, the Independent Data Monitoring Committee did review accumulating serious adverse event data by study arm on one occasion (3 months into study follow-up), at which point they recommended continuation of the trial.

### PATIENT AND PUBLIC INVOLVEMENT

Three patient and public involvement representatives were involved in development of the research questions and the choice of outcome measures specified in the study protocol. One of them also led on development and implementation of strategies to maximise participant recruitment.

## RESULTS

### PARTICIPANTS

A total of 17700 participants in the COVIDENCE UK cohort study were assessed for eligibility to take part in the CORONAVIT trial in October 2020: 6200 of 6470 participants classified as eligible based on their responses to study questionnaires were randomly selected for invitation to the trial, and randomly assigned to higher-dose vs. lower-dose vs. no offer groups (Fig. 1). Table 1 shows baseline characteristics of the trial participants by study arm. Median age was 60.2 years, 4156/6200 (67.0%) were female and 154/6200 (2.5%) had received one or more doses of COVID-19 vaccine. Among participants whose baseline vitamin D status was tested, mean 25(OH)D concentration was 39.7 nmol/L (s.d. 14.5), and 2674/2745 (97.4%) had 25(OH)D concentrations <75 nmol/L. Characteristics were balanced between the three groups. Of 3100 participants randomised to the higher- or lower-dose offer groups, 2958 (95.4%) consented to receive a postal 25(OH)D test, and 2674 (86.3%) had blood 25(OH)D concentrations <75 nmol/L with provision of study supplements (1346 vs. 1328 supplied with 3200 IU vs. 800 IU capsules, respectively). Follow-up was for 6.0 months, from December 2020 to June 2021; by the end of this period, 5523/6200 (89.1%) of participants had received one or more doses of a COVID-19 vaccine (Table S3, Supplementary Appendix). Self-reported adherence to study supplements among participants randomised to either intervention arm was good, with 90.9% of participants reporting that they took study supplements at least 6 times per week (Table S4, Supplementary Appendix). Questionnaire and linkage data availability was also good, with 5979/6200 (96.4%) randomised participants contributing data to the ITT analysis. In the subset of participants included in the ITT analysis for whom measures of end-study vitamin D status was available, mean 25(OH)D concentrations were significantly elevated in the higher-dose vs. no offer group (102.9 vs. 66.6 nmol/L respectively; mean difference 36.3 nmol/L, 95% CI 32.9 to 39.6 nmol/L), and in the lower-dose vs. no offer group (79.4 vs. 66.6 nmol/L respectively; mean difference 12.7 nmol/L, 95% CI 9.8 to 15.6 nmol/L; Table 2, Fig. 2). Among those included in the sensitivity analysis (i.e. excluding non-adherent participants randomised to intervention, and participants in the no offer arm who took vitamin D supplements), mean differences in end-study 25(OH)D concentrations between intervention vs. no offer arms were greater (for higher-dose vs. no offer group, 49.7 nmol/L, 95% CI 45.1 to 54.2 nmol/L; for lower-dose vs. no offer group, 25.8 nmol/L (95% CI 22.0 to 29.5 nmol/L; Table 3, Fig. 2).

**Figure 1:**
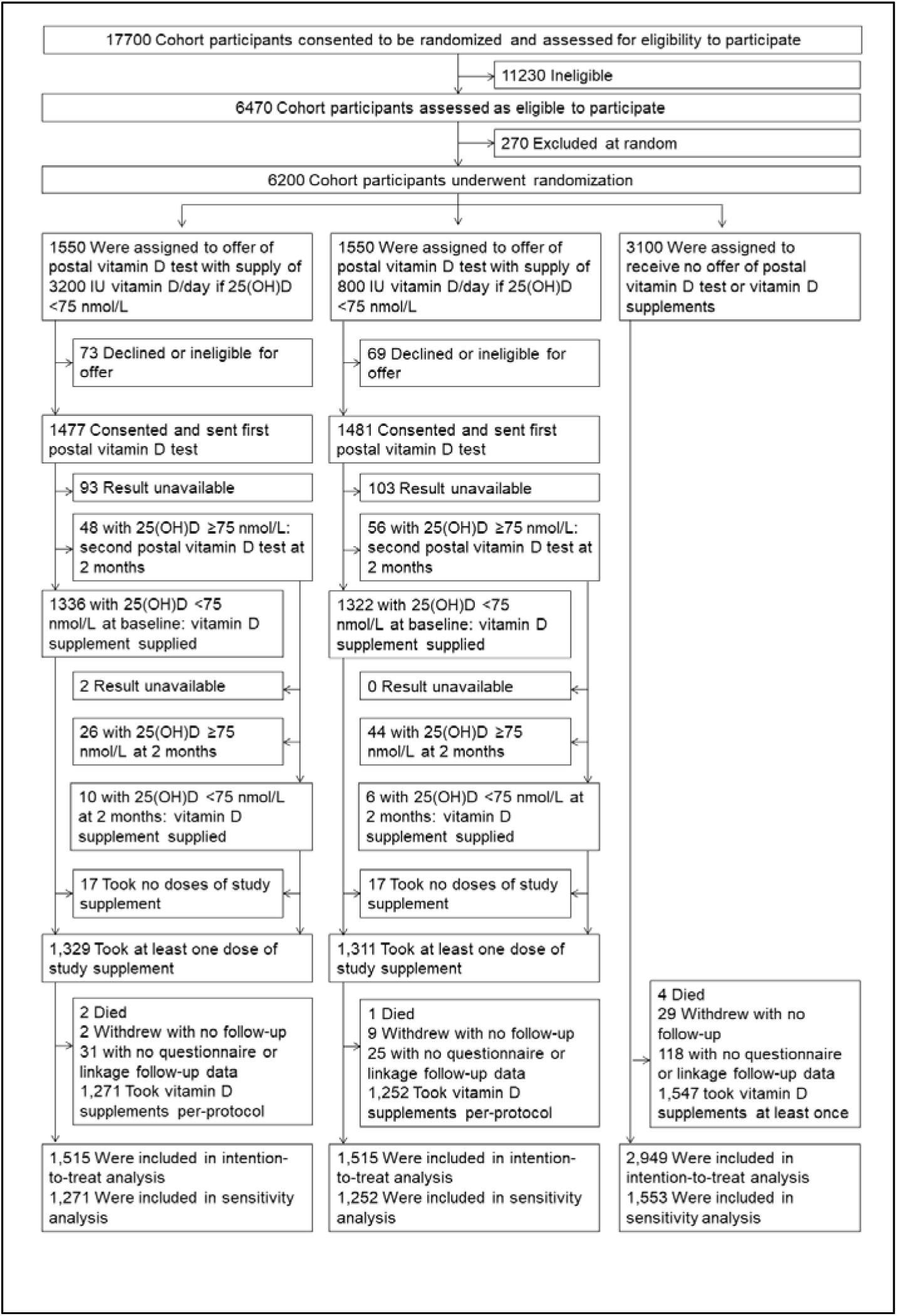
Screening, Randomisation, and Follow-up of the Participants

**Figure 2:**
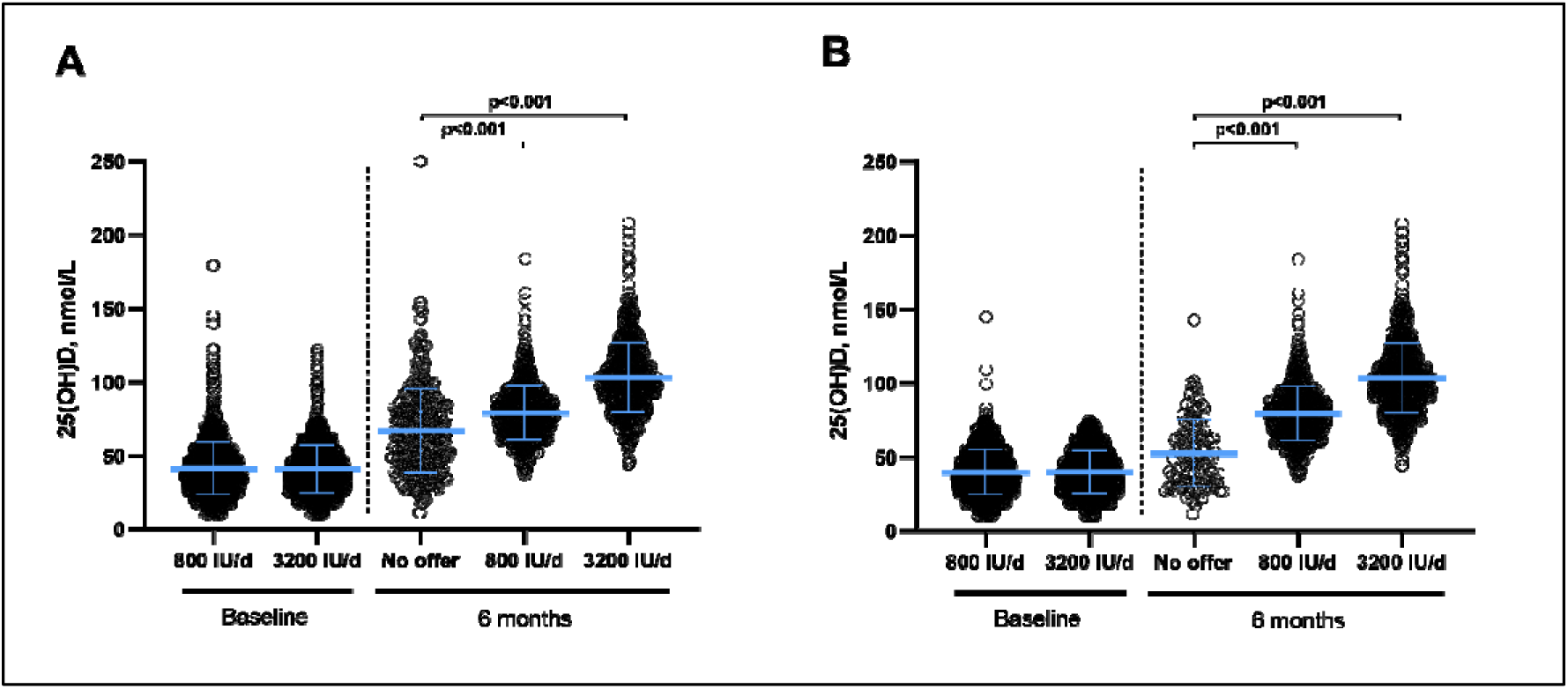
25-hydroxyvitamin D (25[OH]D) concentrations by time-point and allocation, A for participants included in the intention-to-treat analysis, and B, for participants included in the sensitivity analysis excluding data from participants randomised to either intervention arm who reported that they took vitamin D capsules ‘less than half the time’ and those randomised to the no offer arm who reported any intake of supplemental vitamin D during follow-up. Bars show mean and standard deviation for each group. P values are from unpaired Student’s t-tests.

**Table 1:** Baseline Characteristics of the Participants

|  |  | Overall (n=6200) | 3200 IU/day offer (n=1550) | 800 IU/day offer (n=1550) | No offer (n=3100) |
| --- | --- | --- | --- | --- | --- |
| Age | Median age, years (IQR) | 60.2 (49.8 – 67.8) | 60.7 (50.2 – 68.5) | 59.8 (50.3 – 67.4) | 60.8 (49.9 – 68.2) |
|  | Age range, years | 16.1 – 89.8 | 16.4 – 88.6 | 16.5 – 88.2 | 16.1 – 89.8 |
| Sex, N (%) | Male | 2044 (33.0) | 506 (32.6) | 498 (32.1) | 1040 (32.5) |
|  | Female | 4156 (67.0) | 1044 (67.4) | 1052 (67.9) | 2060 (64.4) |
| Ethnicity, N (%) | White | 5867 (94.6) | 1469 (94.8) | 1473 (95.0) | 2925 (94.4) |
|  | Asian/Asian British | 142 (2.3) | 31 (2.0) | 32 (2.1) | 79 (2.5) |
|  | Black/African/Caribbean/Black British | 33 (0.5) | 11 (0.7) | 10 (0.6) | 12 (0.4) |
|  | Mixed/Multiple/Other | 154 (2.5) | 37 (2.4) | 34 (2.2) | 83 (2.6) |
| Country of residence, N (%) | England | 5515 (89.0) | 1374 (88.6) | 1384 (89.3) | 2757 (86.2) |
|  | Northern Ireland | 123 (2.0) | 29 (1.9) | 33 (2.1) | 61 (1.9) |
|  | Scotland | 340 (5.5) | 97 (6.3) | 74 (4.8) | 169 (5.3) |
|  | Wales | 222 (3.6) | 50 (3.2) | 59 (3.8) | 113 (3.5) |
| Highest educational level attained, N (%) | Primary/Secondary | 52 (0.8) | 17 (1.1) | 12 (0.8) | 23 (0.7) |
|  | Higher/Further (A levels) | 924 (14.9) | 215 (13.9) | 233 (15.0) | 476 (14.9) |
|  | College | 2740 (44.2) | 674 (43.5) | 700 (45.2) | 1366 (42.7) |
|  | Post-graduate | 1817 (29.3) | 473 (30.5) | 459 (29.6) | 885 (27.7) |
| Occupational status, N (%) | Employed, health or social care worker | 566 (9.1) | 149 (9.6) | 147 (9.5) | 270 (8.7) |
|  | Employed, other frontline worker | 755 (12.2) | 182 (11.8) | 192 (12.4) | 381 (12.3) |
|  | Employed, non-frontline worker | 1,406 (22.7) | 336 (21.7) | 348 (22.5) | 722 (23.3) |
|  | Self-employed | 564 (9.1) | 154 (9.9) | 144 (9.3) | 266 (8.3) |
|  | Retired | 2504 (40.4) | 635 (41.0) | 606 (39.1) | 1263 (39.5) |
|  | Furloughed | 141 (2.3) | 32 (2.1) | 43 (2.8) | 66 (2.1) |
|  | Unemployed | 126 (2.0) | 34 (2.2) | 42 (2.7) | 50 (1.6) |
|  | Student | 150 (2.4) | 33 (2.1) | 34 (2.2) | 83 (2.6) |
|  | Other | 147 (2.4) | 35 (2.3) | 36 (2.3) | 76 (2.4) |
| Body mass index, kg/m<sup>2</sup>, N (%) | <25 | 2903 (46.8) | 739 (47.7) | 724 (46.7) | 1440 (45.0) |
|  | 25-30 | 2036 (32.8) | 514 (33.2) | 496 (32.0) | 1026 (32.1) |
|  | >30 | 1249 (20.1) | 297 (19.2) | 322 (20.8) | 630 (19.7) |
| Medically diagnosed disease, N (%) | Hypertension | 227 (3.7) | 53 (3.4) | 54 (3.5) | 120 (3.8) |
|  | Diabetes | 259 (4.2) | 81 (5.2) | 56 (3.6) | 122 (3.8) |
|  | Heart disease | 1207 (19.5) | 319 (20.6) | 298 (19.2) | 590 (18.4) |
|  | Asthma | 946 (15.3) | 215 (13.9) | 265 (17.1) | 466 (14.6) |
|  | COPD | 114 (1.8) | 26 (1.7) | 27 (1.7) | 61 (1.9) |
| Tobacco smoking history, N (%) | Never-smoker | 3460 (55.8) | 864 (55.7) | 887 (57.2) | 1709 (53.4) |
|  | Ex-smoker | 2346 (37.8) | 583 (37.6) | 553 (35.7) | 1210 (37.8) |
|  | Current smoker | 393 (6.3) | 103 (6.6) | 109 (7.0) | 181 (5.7) |
| Alcohol consumption/week, units, N (%) | None | 1651 (26.6) | 383 (24.7) | 411 (26.5) | 857 (26.8) |
|  | 1-14 | 3403 (54.9) | 873 (56.3) | 865 (55.8) | 1665 (52.0) |
|  | ≥15 | 1145 (18.5) | 294 (19.0) | 274 (17.7) | 577 (18.0) |
| COVID-19 vaccination status | Unvaccinated | 5774 (93.1) | 1483 (95.7) | 1465 (94.5) | 2826 (91.2) |
|  | Partially vaccinated | 55 (0.9) | 9 (0.6) | 19 (1.2) | 27 (0.9) |
|  | Fully vaccinated | 22 (0.4) | 6 (0.4) | 5 (0.3) | 11 (0.4) |
|  | Not known / missing data | 349 (5.6) | 52 (3.4) | 61 (3.9) | 236 (7.6) |
| Mean 25(OH)D, nmol/L (s.d.) [range]<sup>(1)</sup> |  | --<sup>(2)</sup> | 40.9 (16.4) [10.3-122.0] | 41.5 (18.0) [10.3-179.6] | --<sup>(2)</sup> |
| 25(OH)D category, nmol/L, N (%)<sup>(1)</sup> | <25.0 | --<sup>(2)</sup> | 216 (13.9) | 232 (15.0) | --<sup>(2)</sup> |
|  | 25.0-49.9 | --<sup>(2)</sup> | 797 (51.4) | 759 (49.0) | --<sup>(2)</sup> |
|  | 50-74.9 | --<sup>(2)</sup> | 333 (21.5) | 337 (21.7) | --<sup>(2)</sup> |
|  | ≥75.0 | --<sup>(2)</sup> | 28 (1.8) | 43 (2.7) | --<sup>(2)</sup> |
|  | Not determined | --<sup>(2)</sup> | 176 (11.4) | 179 (11.6) | 3100 (100.0) |

Abbreviations: IQR, inter-quartile range; s.d., standard deviation; 25(OH)D, 25-hydroxyvitamin D.
(1) Missing values: 25(OH)D concentration missing for 189 participants in 3200 IU/day arm and 198 participants in 800 IU/day arm. (2) Baseline 25(OH)D not determined for participants randomised to the 'no offer' arm

**Table 2.**
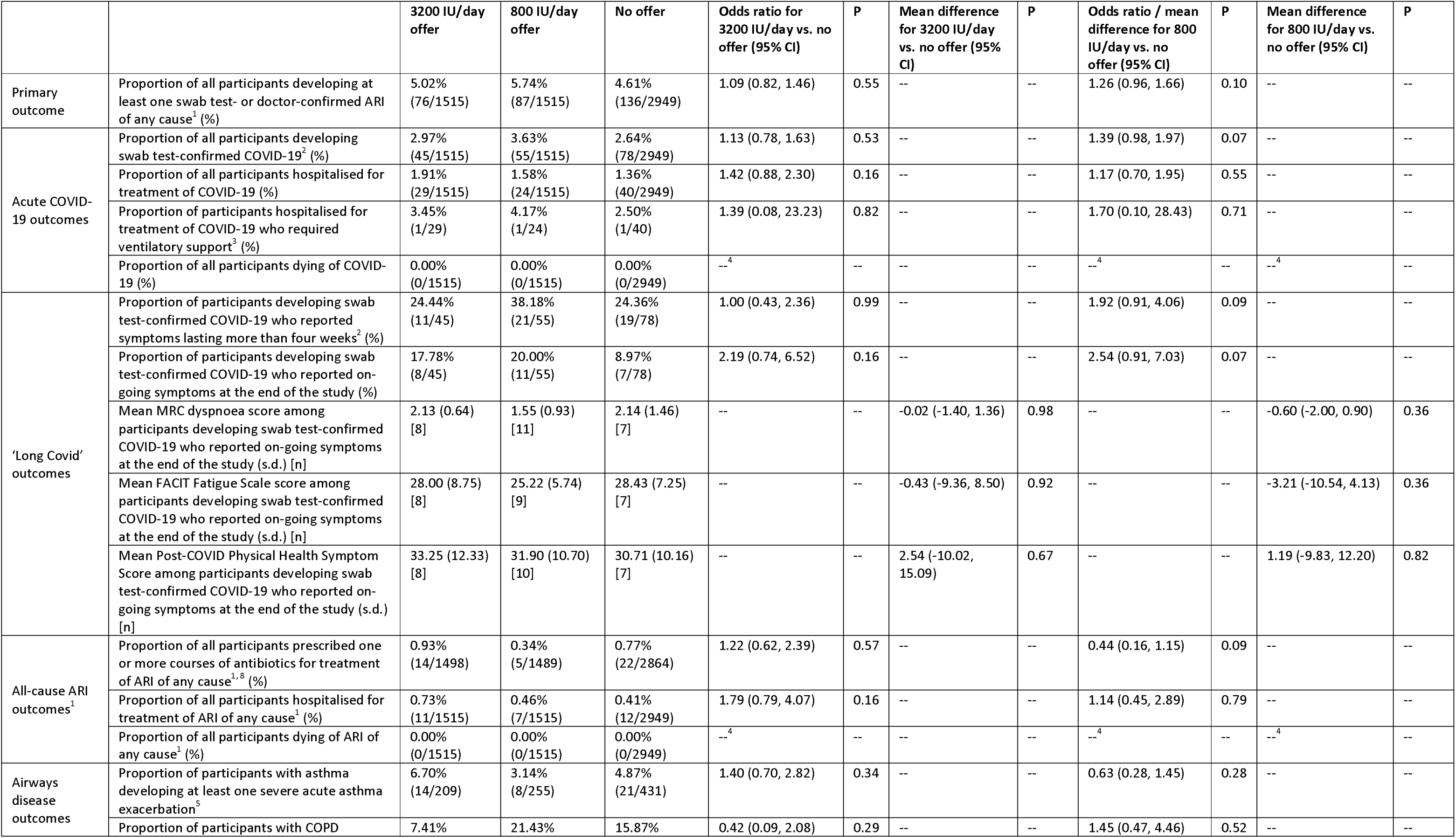
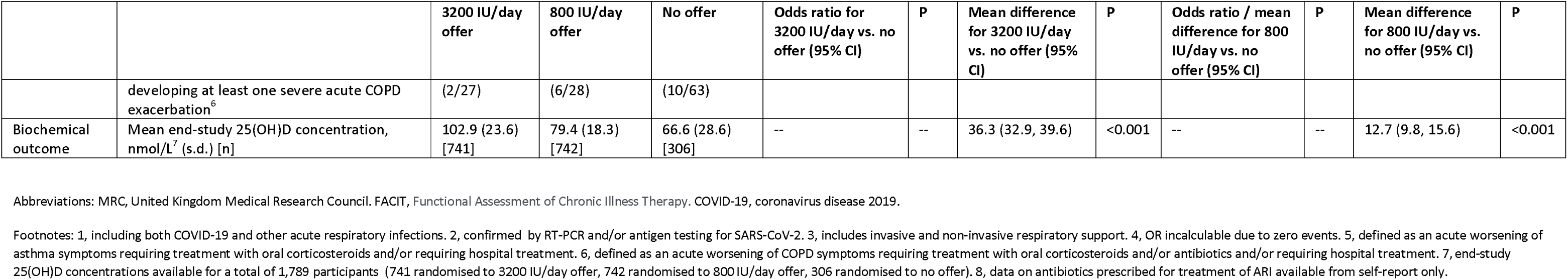
Primary and Secondary Outcomes, by Allocation: Intention-to-Treat Analysis

**Table 3.** Primary and Secondary Outcomes, by Allocation: Sensitivity Analysis^1^

|  |  | 3200 IU/day<br>offer | 800 IU/day<br>offer | No offer | Odds ratio for<br>3200 IU/day<br>vs. no offer<br>(95% CI) | P | Mean difference<br>for 3200 IU/day<br>vs. no offer (95%<br>CI) | P | Odds ratio / mean<br>difference for 800<br>IU/day vs. no offer<br>(95% CI) | P | Mean difference<br>for 800 IU/day vs.<br>no offer (95% CI) | P |
| --- | --- | --- | --- | --- | --- | --- | --- | --- | --- | --- | --- | --- |
| Primary<br>outcome | Proportion of all participants developing at least one swab test- or doctor-confirmed ARI of any cause<sup>2</sup> (%) | 4.33%<br>(55/1269) | 5.31%<br>(66/1243) | 4.43%<br>(59/1331) | 0.98 (0.67, 1.42) | 0.90 | -- | -- | 1.21 (0.84, 1.73) | 0.30 | -- | -- |
| Acute COVID-19 outcomes | Proportion of all participants developing swab test-confirmed COVID-19<sup>3</sup> (%) | 2.52%<br>(32/1269) | 3.14%<br>(39/1243) | 2.55%<br>(34/1331) | 0.99 (0.61, 1.61) | 0.96 | -- | -- | 1.24 (0.77, 1.97) | 0.37 | -- | -- |
|  | Proportion of all participants hospitalised for treatment of COVID-19 (%) | 1.65%<br>(21/1269) | 1.37%<br>(17/1243) | 1.50%<br>(20/1331) | 1.10 (0.59, 2.04) | 0.31 | -- | -- | 0.91 (0.47, 1.74) | 0.77 | -- | -- |
|  | Proportion of participants hospitalised for treatment of COVID-19 who required ventilatory support<sup>4</sup> (%) | 4.76%<br>(1/21) | 0.00%<br>(0/17) | 5.00%<br>(1/20) | 0.95 (0.55, 16.29) | 0.97 | -- | -- | --<sup>4</sup> | -- | -- | -- |
|  | Proportion of all participants dying of COVID-19 (%) | 0.00%<br>(0/1269) | 0.00%<br>(0/1243) | 0.00%<br>(0/1331) | --<sup>4</sup> | -- | -- | -- | --<sup>4</sup> | -- | -- | -- |
| 'Long Covid' outcomes | Proportion of participants developing swab test-confirmed COVID-19 who reported symptoms lasting more than four weeks<sup>3</sup> (%) | 31.25%<br>(10/32) | 35.90%<br>(14/39) | 26.47%<br>(9/34) | 1.26 (0.43, 3.67) | 0.67 | -- | -- | 1.56 (0.57, 4.25) | 0.39 | -- | -- |
|  | Proportion of participants developing swab test-confirmed COVID-19 who reported on-going symptoms at the end of the study (%) | 18.75%<br>(6/32) | 20.51%<br>(8/39) | 5.88%<br>(2/34) | 3.69 (0.69, 19.85) | 0.13 | -- | -- | 4.13 (0.81, 21.00) | 0.09 | -- | -- |
|  | Mean MRC dyspnoea score among participants developing swab test-confirmed COVID-19 who reported on-going symptoms at the end of the study (s.d.) [n] | 2.29 (0.49)<br>[7] | 1.29 (0.76)<br>[7] | 1.00<sup>6</sup><br>[1] | -- | -- | 1.29<sup>7</sup> | -- | -- | -- | 0.29<sup>7</sup> | -- |
|  | Mean FACIT Fatigue Scale score among participants developing swab test-confirmed COVID-19 who reported on-going symptoms at the end of the study (s.d.) [n] | 25.29 (4.53)<br>[7] | 23.00 (4.69)<br>[6] | 34.00<sup>6</sup><br>[1] | -- | -- | -8.71<sup>7</sup> | -- | -- | -- | -11.00<sup>7</sup> | -- |
|  | Mean Post-COVID Physical Health Symptom Score among participants developing swab test-confirmed COVID-19 who reported on-going symptoms at the end of the study (s.d.) [n] | 30.57 (10.50)<br>[7] | 30.50 (11.33)<br>[6] | 29.00<sup>6</sup><br>[1] | -- | -- | 1.57<sup>7</sup> | -- | -- | -- | 1.50<sup>7</sup> | -- |
| All-cause ARI outcomes<sup>1</sup> | Proportion of all participants prescribed one or more courses of antibiotics for treatment of ARI of any cause<sup>2,11</sup> (%) | 0.55%<br>(7/1269) | 0.32%<br>(4/1243) | 0.98%<br>(13/1331) | 0.56 (0.22, 1.41) | 0.22 | -- | -- | 0.33 (0.11, 1.00) | 0.05 | -- | -- |
|  | Proportion of all participants hospitalised for treatment of ARI of any cause<sup>1</sup> (%) | 0.79%<br>(10/1269) | 0.24%<br>(3/1243) | 0.45%<br>(6/1331) | 1.75 (0.64, 4.84) | 0.28 | -- | -- | 0.53 (0.13, 2.14) | 0.38 | -- | -- |
|  | Proportion of all participants dying of ARI of any cause<sup>2</sup> (%) | 0.00%<br>(0/1269) | 0.00%<br>(0/1243) | 0.00%<br>(0/1331) | --<sup>5</sup> | -- | -- | -- | --<sup>5</sup> | -- | -- | -- |
| Airways disease outcomes | Proportion of participants with asthma developing at least one severe acute asthma exacerbation<sup>8</sup> | 4.73%<br>(8/169) | 3.33%<br>(7/210) | 6.11%<br>(11/180) | 0.76 (0.30, 1.95) | 0.57 | -- | -- | 0.53 (0.20, 1.40) | 0.20 | -- | -- |
|  | Proportion of participants with COPD developing at least one severe acute COPD | 4.55%<br>(1/22) | 25.00%<br>(6/24) | 17.86%<br>(5/28) | 0.22 (0.02, 2.03) | 0.18 | -- | -- | 1.53 (0.40, 5.84) | 0.53 | -- | -- |

|  |  | **3200 IU/day offer** | **800 IU/day offer** | **No offer** | **Odds ratio for 3200 IU/day vs. no offer (95% CI)** | **P** | **Mean difference for 3200 IU/day vs. no offer (95% CI)** | **P** | **Odds ratio / mean difference for 800 IU/day vs. no offer (95% CI)** | **P** | **Mean difference for 800 IU/day vs. no offer (95% CI)** | **P** |
| --- | --- | --- | --- | --- | --- | --- | --- | --- | --- | --- | --- | --- |
|  | exacerbation<sup>9</sup> |  |  |  |  |  |  |  |  |  |  |  |
| **Biochemical outcome** | Mean end-study 25(OH)D concentration, nmol/L<sup>10</sup> (s.d.) [n] | 103.4 (23.3) [729] | 79.5 (18.3) [736] | 53.7 (23.1) [116] | -- | -- | 49.7 (45.1, 54.2) | <0.001 | -- | -- | 25.8 (22.0, 29.5) | <0.001 |

Abbreviations: MRC, United Kingdom Medical Research Council; FACIT, Functional Assessment of Chronic Illness Therapy; COVID-19, coronavirus disease 2019.
Footnotes: 1, this analysis excludes data from participants randomised to either intervention arm who reported that they took vitamin D capsules 'less than half the time' as well as those randomised to the no offer arm who reported any intake of supplemental vitamin D during follow-up. 2, including both COVID-19 and other acute respiratory infections. 3, confirmed by RT-PCR and/or antigen testing for SARS-CoV-2. 4, includes invasive and non-invasive respiratory support. 5, OR in calculable due to zero events. 6, s.d. in calculable as single participant with outcome in no offer arm. 7, 95% CI in calculable as single participant with outcome in no offer arm. 8, defined as an acute worsening of asthma symptoms requiring treatment with oral corticosteroids and/or requiring hospital treatment. 9, defined as an acute worsening of COPD symptoms requiring treatment with oral corticosteroids and/or antibiotics and/or requiring hospital treatment. 10, end-study 25(OH)D concentrations available for a total of 1,789 participants (741 randomised to 3200 IU/day offer, 742 randomised to 800 IU/day offer, 306 randomised to no offer). 11, data on antibiotics prescribed for treatment of ARI available from self-report only.

### PRIMARY AND SECONDARY OUTCOMES

The primary end point of at least one episode of swab test- or doctor-confirmed ARI occurred in 290 participants, with no statistically significant difference in proportions experiencing such an event in either offer group vs. the no offer group (for higher-dose vs. no offer: 76/1515 (5.0%) vs. 136/2949 (4.6%), respectively; OR 1.09; 95% CI 0.82 to 1.46, P=0.55; for lower-dose vs. no offer: 87/1515 (5.7%) vs. 136/2949 (4.6%), respectively; OR 1.26; 95% CI 0.96 to 1.66, P=0.10; Table 2).

No statistically significant differences in outcomes relating to incidence or severity of acute COVID-19, prolonged symptoms of COVID-19 were seen between those randomised to either offer vs. no offer (Table 2). We also found no evidence to suggest that allocation to either offer vs. no offer influenced prescription of antibiotics for ARI treatment, hospitalisation or death from all-cause ARI, or incidence of acute exacerbations of asthma or COPD (Table 2).

### SUB-GROUP ANALYSES

Sub-group analysis revealed no evidence to suggest that COVID-19 vaccination modified the effect of allocation on incidence of COVID-19 or prolonged COVID-19 symptoms (Table S5, Supplementary Appendix). Planned sub-group analysis by baseline vitamin D status was not conducted, as the range and distribution of imputed 25(OH)D concentrations in the ‘no offer’ arm at baseline did not match those of measured 25(OH)D concentrations in participants randomised to either intervention arm (Figure S1, Supplementary Appendix), calling the validity of the imputation into question. Exploratory survival analysis to determine effects of allocation before vs. after administration of a first dose of COVID-19 vaccine did not show protective effects of the intervention during either phase of follow-up (for lower-dose vs. no offer, pre-vaccination phase, hazard ratio [HR] 1.37, 95% CI 0.90 to 2.07, P=0.14; post-vaccination phase, HR 1.68, 95% CI 0.87 to 3.22, P=0.12; for higher-dose vs. no offer, pre-vaccination phase, HR 0.93, 95% CI 0.58 to 1.49, P=0.73; post-vaccination phase, HR 1.57, 95% CI 0.81 to 3.05, P=0.18; Figure S2, Supplementary Appendix).

### SENSITIVITY ANALYSES

Of 3100 people randomised to the no offer group, 1547 (49.9%) reported that they took supplemental vitamin D on at least one occasion during the study, while 2523/2674 (94.4%) participants supplied with study supplements reported that they took them more than half the time. Results of sensitivity analyses excluding the former group and including the latter (Table 3) were not materially different to those yielded by ITT analyses (Table 2). A post hoc exploratory analysis comparing mean end-study 25(OH)D concentrations of control arm participants who contributed data to the ITT analysis but not to the sensitivity analysis (i.e. participants randomised to no offer who reported use of off-trial vitamin D supplements) vs. those who contributed data to both ITT & sensitivity analyses (i.e. participants randomised to no offer who reported no use of off-trial vitamin D supplements) revealed mean end-trial 25(OH)D concentrations to be higher in the former group vs. the latter (74.5 vs 53.7 nmol/L; 95% CI for difference, 14.6 to 27.0 nmol/L, p<0.001). Another post hoc exploratory sensitivity analysis evaluating the effect of allocation among participants who completed all six follow-up questionnaires yielded null results (OR for higher-dose vs. no offer 1.01, 95% CI 0.73 to 1.39, P=0.96; OR for lower-dose vs. no offer 1.16, 95% CI 0.85 to 1.56, P=0.35).

### ADVERSE EVENTS

7 participants (2 vs. 1 vs. 4 allocated to higher-dose vs. lower-dose vs. no offer groups, respectively) died during the study, and 313 (85 vs. 85 vs. 143 allocated to higher-dose vs. lower-dose vs. no offer groups) experienced one or more non-fatal serious adverse events (Table S6, Supplementary Appendix). Causes of these events are presented in Table S7, Supplementary Appendix: none was adjudged to be related to administration of study supplements. Four participants in the higher dose offer group developed hypercalcemia (serum corrected calcium >2.65 mmol/L): study supplements were discontinued, and the hypercalcemia and symptoms resolved. One participant in the no offer group was found to have asymptomatic hypervitaminosis D (25[OH]D 250 nmol/L) at 6-month follow-up, after taking a non-study vitamin D supplement at a dose of 4000 IU/day. One participant in the higher dose offer group was hospitalised on two occasions with renal colic due to nephrolithiasis. A total of 47 non-severe adverse events led to discontinuation of study supplements (24 vs. 23 in higher- vs. lower-dose offer groups, respectively: Table S8, Supplementary Appendix).

## DISCUSSION

We present results of the first phase 3 RCT to evaluate the effectiveness of a test-and-treat approach to correction of sub-optimal vitamin D status for prevention of ARIs. It is also the first clinical trial to investigate whether vitamin D supplementation reduces risk of COVID-19. Among participants randomised to receive an offer of vitamin D testing, uptake of this intervention was good, prevalence of 25(OH)D concentrations <75 nmol/L was high, and end-study 25(OH)D concentrations were elevated when compared to those who were randomised to no such offer, providing objective evidence of a high level of adherence. However, no statistically significant effect of either dose was seen on the primary outcome of incident doctor- or swab test-confirmed ARI, or on the major secondary outcome of incident swab test-confirmed COVID-19. Oral vitamin D supplementation was safe and well-tolerated at both doses investigated: incidence of adverse events was balanced between arms, and no serious adverse event was attributed to study supplements.

The design of our study was informed by findings from a recent meta-analysis, suggesting that protective effects of vitamin D against ARI might be strongest when daily doses of 400-1000 IU were given for up to one year.^22^ The results from the current study do not support the hypothesis that such regimens offer protection against ARI, and are consistent with those of several other recent phase 3 trials of vitamin D supplementation that have reported no effect of vitamin D supplementation on risk of ARIs.^20^ ^21^ ^38^ The null result for the major secondary outcome of incident COVID-19 in this trial is consistent with our finding of no independent association between intake of supplemental vitamin D and risk of COVID-19 in a prospective observational study undertaken in this cohort prior to initiation of this trial,^12^ as well as null results from a Mendelian randomisation study that tested for associations between genetically predicted 25(OH)D concentrations and susceptibility to COVID-19.^39^ However, it contrasts with recent findings from a Phase 2 RCT conducted in Mexican health care workers, which reported a strong protective effect of a daily dose of 4,000 IU vitamin D.^23^ Contrasting findings may relate to the fact that participants in the Mexican trial were SARS-CoV-2 vaccine-naïve, and/or the relatively short duration of follow-up (one month). The former hypothesis is not supported by results from sub-group analysis of the current study, showing no effect of vitamin D on incident COVID-19 either before or after SARS-CoV-2 vaccination.

Our study has several strengths. In contrast to recent large clinical trials of vitamin D supplementation for the prevention of ARIs,^21^ ^38^ our study population had a very high prevalence of sub-optimal vitamin D status at baseline, with 97.4% of those tested having 25(OH)D concentrations <75 nmol/L. We investigated two dosing regimens utilising daily dosing (thereby avoiding large and unphysiological fluctuations in 25[OH]D that are seen with administration of intermittent bolus dosing),^40^ and there was good adherence (evidenced by self-report and by significant differences in end-study 25[OH]D concentrations between arms). The trial within cohort design allowed a rapid and efficient evaluation of a pragmatic approach to boosting vitamin D status in the general population to provide a timely answer to a pressing global public health question. Linkage with routinely collected data from medical records allowed comprehensive capture of outcomes in those who did not complete study questionnaires, allowing us to minimise loss to follow-up and to capture important events that precluded questionnaire completion such as severe illness and death. The trial was initiated prior to widespread roll-out of COVID-19 vaccination, and follow-up coincided with the ‘second wave’ of COVID-19 in the UK: both factors contributed to the appreciable number of COVID-19 cases that arose, which allowed for potential effects of vitamin D on prevention of this specific cause of ARI to be investigated. Other strengths include a rigorous case definition for the primary outcome that required objective confirmation of ARI (as opposed to self-report of symptoms), and use of an externally accredited laboratory to measure vitamin D status using liquid chromatography-tandem mass spectrometry, which is the gold standard assay for this determination.

Our study also has limitations. Provision of supplements to participants randomised to intervention was contingent on demonstrating inadequate vitamin D status: thus, a subset (13.7%) of participants randomised to intervention did not receive study supplements. On the other hand, another subset (49.9%) of participants randomised to no offer took a vitamin D supplement on one or more occasions during follow-up.

This may have led to increases in 25(OH)D concentrations in the no offer arm over the course of the study, although seasonal effects (sampling in June vs. December) will also have contributed. Together, these factors could have diluted any effect of vitamin D in the primary ITT analysis. We sought to overcome this by conducting a sensitivity analysis, which included only those randomised to offer vs. no offer who did vs. did not take supplemental vitamin D, respectively. The fact that this analysis showed no effect of vitamin D supplementation on all outcomes investigated, despite the larger differences in end-study 25(OH)D concentrations between intervention vs. no offer arms seen for this analysis vs. the ITT analysis (Fig. 2), provides some reassurance that the null result yielded by the ITT analysis is valid. Ultimately, however, this trial was designed to investigate the effectiveness of a pragmatic ‘test-and-treat’ approach to boosting population vitamin D status, rather than biologic efficacy of vitamin D to prevent ARIs, and our findings should be interpreted accordingly: specifically, we highlight that they are not inconsistent with findings from meta-analyses of placebo-controlled trials of vitamin D to prevent ARI,^22^ ^41^ which better address questions of efficacy. The open-label design may have introduced ascertainment bias by influencing the likelihood of participants completing follow-up questionnaires. This potential problem was off-set by use of medical record linkage, which allowed us to capture outcomes in those did not complete all follow-up questionnaires. Moreover, a post hoc sensivitity analysis restricted to participants who completed all follow-up questionnaires yielded null results, consistent with those of the primary analysis. The proportion of those randomised to ‘no offer’ who experienced the primary outcome (4.6%) was lower than the 20% anticipated in the sample size calculation, possibly reflecting the impact of public health measures to control transmission of SARS-CoV-2 (such as lockdowns, social distancing and mask wearing) on incidence of other ARIs.^42^ Alternatively, it might reflect a reduction in consultations for ARI arising as a result of participants’ reluctance to attend a doctor’s surgery with ARI symptoms, as face-to-face consultations were actively discouraged during the pandemic, and there was a reticence among the general public to over-burden a health service that was already over-stretched.^43^ ^44^ This could have compromised power; however, the lower bounds for the 95% CIs of ORs relating to the effect of higher- or lower-dose offers on our primary outcome (0.82 and 0.96, respectively) effectively rule out relative reductions in odds of ARI of more than 18% and 4%, respectively. Arguably, effects of this size or less are unlikely to be considered of sufficient magnitude to implement the study intervention for the purpose of ARI prevention. Incidence of some secondary outcomes, including hospitalisation for ARI, was low: we therefore lacked power to detect an effect of the intervention on severity of COVID-19 and other ARIs. Prevalence of profound vitamin D deficiency (25[OH]D <25 nmol/L) at baseline was also low, and we therefore lacked power to detect an effect of the intervention in this group, who may be more likely to derive clinical benefit from vitamin D replacement than those with higher baseline 25(OH)D concentrations.^45^ Finally, we acknowledge that men, ethnic minorities and people with lower educational attainment were relatively under- represented among study participants vs. the general population, which may have compromised the generalisability of our findings. However, we also highlight that other groups at increased risk of severe COVID-19 were over-represented among trial participants, of whom 35.6% were aged ≥65 years (compared to 18.3% of the UK population)^46^ and 19.5% had heart disease (compared to 3-4% of the UK population).^47^ This, together with the fact that many participants were unvaccinated or partially vaccinated during follow-up, may explain why a relatively high proportion of participants with COVID-19 required hospitalisation.

In conclusion, we report that implementation of a test-and-treat approach to correcting sub-optimal vitamin D status in the U.K. population was safe and effective in boosting 25(OH)D concentrations of adults with baseline concentrations <75 nmol/L. However, this was not associated with protection against all-cause ARI or COVID-19.

## DATA SHARING STATEMENT

Anonymised patient-level data will be made available on reasonable request to, subject to the terms of Research Ethics Committee and Sponsor approval.

## ETHICS APPROVAL

The study was approved by the Queens Square Research Ethics Committee, London, UK (ref 20/HRA/5095) and all participants gave informed consent before taking part.

## TRANSPARENCY STATEMENT

ARM affirms that the manuscript is an honest, accurate, and transparent account of the study results. All outcomes pre-specified in the protocol and the trial registry are reported here, except for those relating to potential effects of vitamin D on SARS-CoV-2 vaccine immunogenicity, which will be reported in a separate manuscript.

## Supporting information

Supplementary Appendix CORONAVIT

## Data Availability

Anonymised data will be provided on reasonable request to subject to the terms of ethical approval and Sponsor requirements.

## ACKNOWLEDGEMENTS

We thank all the people who participated in the trial; members of the Independent Data Monitoring Committee (Prof Irwin Nazareth, University College London [Chair]; Dr Michael Grayling; and Dr Richard Quinton, University of Newcastle upon Tyne); members of the Trial Steering Committee (Prof Paul Lips, Amsterdam University Medical Centre, Amsterdam [Chair]; Dr Gwyneth Davies, University College London; and Dr Anna Bibby, University of Bristol); and Ms. Mujiba Ejaz (N.H.S. Digital), Ms. Caroline Brooks (Swansea University) and Ms. Susan Knight (Public Health Scotland) for assistance with medical record data linkage.

## DISSEMINATION

Trial participants were emailed the following link to a webinar in which results are presented in plain language: https://www.youtube.com/watch?v=QpVl0xlRP0A. The trial report was published on a pre-print server (https://www.medrxiv.org/content/10.1101/2022.03.22.22271707v1), and results attracted the interest of national and international media (e.g. https://inews.co.uk/news/vitamin-d-supplements-do-not-stop-you-catching-covid-or-reduce-symptoms-major-trial-reveals-1536673 , https://www.news-medical.net/news/20220325/Effectiveness-of-a-test-and-treat-approach-for-identification-and-treatment-of-vitamin-D-insufficiency-for-prevention-of-COVID-19.aspx).

## STUDY DESIGN AND AUTHOR CONTRIBUTIONS

ARM, DAJ and CR designed the study, with input from PP, JS, DF, RAL, GAD, FK, CJG, JN, AS, SEF, AGR and SOS. The trial was managed by DAJ, HH, NP, SM, MT and ARM. Laboratory assays were performed by AN, NLB and RG. Data were managed and analysed by DAJ, MG, MT and CO. All the authors vouch for the accuracy and completeness of the data and for the fidelity of the trial to the protocol. ARM wrote the first draft of the paper. All authors contributed to the interpretation of the results, review and approval of the manuscript, and the decision to submit it for publication. There were no agreements concerning confidentiality of the data between the sponsor and the authors or the institutions named in the credit lines.

