## Supplementary Appendix CORONAVIT for "Vitamin D Supplements for Prevention of COVID-19 or other Acute Respiratory Infections: a Phase 3 Randomised Controlled Trial (CORONAVIT)"

This appendix has been provided by the authors to give readers additional information about their work.

**SUPPLEMENTARY APPENDIX**

### Study Personnel

**Chief Investigator:** Prof Adrian R. Martineau (Queen Mary University of London)

**Principal Investigator:** Dr David A. Jolliffe (Queen Mary University of London)

**Trial Steering Committee:** Prof Paul Lips (Chair, VUMC Amsterdam), Dr Anna Bibby (University of Bristol), Dr Gwyneth Davies (University College London)

**Independent Data Monitoring Committee:** Prof Irwin Nazareth (Chair, University College London), Dr Michael Grayling (University of Newcastle upon Tyne), Dr Richard Quinton (University of Newcastle upon Tyne)

**Statisticians:** Dr David A. Jolliffe, Mr Matthew Greenig, Dr Mohammad Talaei, Dr Giulia Vivaldi (Queen Mary University of London).

**Trial Management team:** Dr David A. Jolliffe, Prof Adrian R Martineau, Ms. Natalia Perdek, Ms. Hayley Holt, Ms. Sheena Maltby (Queen Mary University of London)

**Data Management team:** Dr David A. Jolliffe (Queen Mary University of London), Dr Mohammad Talaei (Queen Mary University of London), Mr Chris Orton (Swansea University Medical School)

**Laboratory team:** Dr Nicola Barlow, Ms. Alexa Normandale, Ms. Rajvinder Garcha (Black Country Pathology Services)

Table S1: Questions from monthly follow-up questionnaire capturing self-reported primary outcomes. Caseness for swab test- or doctor-confirmed acute respiratory infection (primary outcome) was ascribed where participants answered (‘Yes’ to Q1.1 AND ‘Yes’ to any of the underlined responses to Q1.3) OR (‘Yes’ to Q2.1 AND ‘Yes’ to Q2.2 AND ‘Yes’ to any of the underlined responses to Q2.4) OR (‘Yes’ to Q2.1 AND ‘Yes’ to Q3.1 AND ‘Yes’ to any of the underlined responses to Q3.3).

| 1. **Self-reported laboratory-confirmed ARI/COVID-19** | |
| --- | --- |
| **1.1** Since you last checked in with us, have you had a nose or throat swab for COVID-19 or any other respiratory virus, or has a result from a previous swab test become newly available?  (This question is about tests to detect the virus itself: they are usually done in somebody who has symptoms, but screening of asymptomatic people can also be done. It’s usually a nose/throat swab, but saliva tests are also becoming available) | • Yes  • No |
| **1.2** On what date did you have this nose / throat swab?  If you are not sure of the exact date, enter the approximate date (DD/MM/YYYY). |  |
| **1.3** What was the result? Click as many as apply. | • Positive for COVID-19 (SARS-CoV-2 coronavirus)  • Positive for influenza virus  • Positive for another respiratory virus  • Negative for all/any viruses tested  • Not known |
| 2. **Self-reported GP-confirmed ARI/COVID-19** | |
| **2.1** Since you last checked in with us, have you experienced any of the following symptoms: cold or flu symptoms, sore throat, persistent cough, loss of smell or taste, fever, fatigue, diarrhoea, abdominal pain or loss of appetite? | • Yes, I have had one or more of these symptoms since completing my last COVIDENCE UK questionnaire  • No, I have not had any of these symptoms since completing my last COVIDENCE UK questionnaire |
| **2.2** Did you have a face-to-face appointment with your GP to discuss these symptoms? | • Yes  • No |
| **2.3** On what date did you have an appointment with your GP (DD/MM/YYYY)? |  |
| **2.4** What did the GP diagnose? Tick as many as apply | • Suspected or proven COVID-19  • Pneumonia  • ‘Flu' (influenza)  • Bronchitis  • Tonsillitis or pharyngitis (sore throat)  • Ear infection (otitis media)  • Common cold  • Another upper respiratory infection  • Another lower respiratory infection  • Something else |
| 3. **Self-reported hospitalisation for ARI/COVID-19** | |
| **3.1** Did you go to hospital for treatment of these symptoms? | • Yes, and I was admitted to hospital (i.e. I spent one or more nights as a hospital in-patient)  • Yes, I attended a hospital accident and emergency department but I was not admitted to hospital (i.e. I went home without spending one or more nights as a hospital in-patient)  • No, I didn’t go to hospital for treatment of these symptoms |
| **3.2** On what date did you attend hospital (DD/MM/YYYY)? |  |
| **3.3** What did the hospital doctors diagnose? Tick as many as apply | • Suspected or proven COVID-19  • Pneumonia  • ‘Flu' (influenza)  • Bronchitis  • Tonsillitis or pharyngitis (sore throat)  • Ear infection (otitis media)  • Common cold  • Another upper respiratory infection  • Another lower respiratory infection  • Something else |

### Table S2: International Classification of Diseases, 10^th^ revision (ICD-10) codes used to capture primary outcomes in databases containing routinely collected virology test results and health data.

Acute Upper Respiratory Infections:

J00	acute nasopharyngitis

J01	acute sinusitis

J010	acute maxillary sinusitis

J011	acute frontal sinusitis

J012	acute ethmoidal sinusitis

J013	acute sphenoial sinusitis

J014	acute pansinusitis

J018	other acute sinusitis

J019	acute sinusitis, unspecified

J02	acute pharyngitis

J020	streptococcal pharyngitis

J028	acute pharyngitis due to other organism

J029	acute pharyngitis, unspecified

J03	acute tonsilitis

J030	streptococcal tonsilitis

J038	acute tonsilitis due to other organism

J039	acute tonsilitis, unspecified

J04	acute laryngitis and tracheitis

J040	acute laryngitis

J041	acute tracheitis

J042	acute laryngotracheitis

J05	acute obstructive laryngitis and epiglottis

J050	acute obstructive laryngitis

J051	acute epiglottis

J06	acute upper respiratory infections of multiple and unspecified sites

J060	acute laryngopharyngitis

J068	other acute upper respiratory infections of multiple sites

J069	acute upper respiratory infection, unspecified

Acute Lower Respiratory Infections:

J12	viral pneumonia

J120	adenoviral pneumonia

J121	RSV pneumonia

J122	parainfluenza virus pneumonia

J123	human metapneumovirus pneumonia

J128	other viral pneumonia

J129	viral pneumonia, unspecified

J13	pneumonia, streptococcus pneumoniae

J14	pneumonia, haemophilus influenzae

J15	bacterial pneumonia, not elsewhere classified

J150	pneumonia, kebsilla pneumoniae

J151	pneumonia, pseudomonas

J152	pneumonia, staphylococcus

J153	pneumonia, streptococcus, group B

J154	pneumonia, other streptococci

J156	pneumonia, other gram-negative bacteria

J157	pneumonia, mycoplasma pneumoniae

J158	pneumonia, other bacterial

J159	pneumonia, bacterial unspecified

J16	pneumonia, other infectious organisms

J160	pneumonia, chlamydial

J168	pneumonia, other specified infectious organism

J17	pneumonia, in diseases classfied elsewhere

J170	pneumonia, bacterial diseases classfied elsewhere

J171	pneumonia, viral diseases classified elsewhere

J172	pneumonia, in mycoses

J173	pneumonia, in parasitic diseases

J178	pneumonia in other diseases classified elsewhere

J18	pneumonia, organism unspecified

J180	bronchopneumonia, unspecified

J181	lobar pneumonia, unspecified

J182	hypostatic pneumonia, unspecified

J188	other pneumonia, organism unspecified

J189	pneumonia, unspecified

J20	acute bronchitis

J200	acute bronchitis, mycoplasma pneumonia

J201	acute bronchitis, haemophilus influenzae

J202	acute bronchitis, streptococcus

J203	acute bronchitis, coxsackievirus

J204	acute bronchitis, parainfluenza virus

J205	acute bronchitis, RSV

J206	acute bronchitis, rhinovirus

J207	acute bronchitis, echovirus

J208	acute bronchitis, other specified organism

J209	acute bronchitis, unspecified

J21	acute bronchiolitis

J210	acute bronchiolitis, RSV

J211	acute bronchiolitis, metapneumovirus

J218	acute bronchiolitis, other specified organisms

J219	acute bronchiolitis, unspecified

J22	unspecified acute lower respiratory infection

Influenza:

J09	influenza, zoonotic or pandemic

J10	influenza, seasonal B/C

J100	influenza (seasonal) with pneumonia

J110	influenza (seasonal) with other respiratory manifestations

J111	influenza with other respiratory manifestations, virus not detected

J112	influenza with gastrointestinal manifestations

J118	influenza wither other manifestations, virus not detected

COVID-19:

U07.1	COVID-19, virus identified

U07.2	COVID-19, virus not identified

### Table S3: End-trial COVID-19 vaccination status by allocation

|  | **3200 IU/day offer (n=1550)** | **800 IU/day offer (n=1550)** | **No offer (n=3100)** |
| --- | --- | --- | --- |
| Unvaccinated | 68 (4.4%) | 69 (4.5%) | 191 (6.2%) |
| Partially vaccinated, N (%)^1^ | 253 (16.3%) | 226 (14.6%) | 514 (16.6%) |
| Fully vaccinated, N (%)^2^ | 1177 (75.9%) | 1194 (77.0%) | 2159 (69.6%) |
| Vaccination status not known, N (%) | 52 (3.4%) | 61 (3.9%) | 236 (7.6%) |

1, of whom 466 (46.9%) received a single dose of ChAdOx1 nCoV-19, 460 (46.3%) received a single dose of BNT162b2 and 67 (6.7%) received a single dose of another COVID-19 vaccine. 2, of whom 2756 (60.8%) received two doses of ChAdOx1 nCoV-19, 1552 (34.3%) received two doses of BNT162b2 and 222 (4.9%) received two doses of mixed or other COVID-19 vaccines.

### Table S4: Self-reported frequency of study capsule intake among participants randomised to intervention arms of the trial who were sent study capsules, as reported in the study adherence questionnaire, sent on 31^st^ March 2021.

|  | **3200 IU/day arm (n=1339)** | **800 IU/day arm (n=1314)** |
| --- | --- | --- |
| Every day, N (%) | 1065 (79.5) | 1052 (80.1) |
| Almost every day (6-7 times per week, on average), N (%) | 143 (10.7) | 151 (11.5) |
| Most days (4-5 times per week, on average), N (%) | 45 (3.4) | 40 (3.0) |
| Less than half the time (1-3 times per week, on average), N (%) | 10 (0.7) | 9 (0.7) |
| Less often than once per week, on average, N (%) | 0 (0) | 0 (0) |
| Not taking them at all, N (%) | 0 (0) | 0 (0) |
| Did not respond, N (%) | 76 (5.7) | 62 (4.7) |

### Table S5: COVID-19 outcomes by allocation and COVID-19 vaccination status at the end of the trial

| **Outcome** | **Sub-group** | **3200 IU/day offer** | **800 IU/day offer** | **No offer** | **Odds ratio for 3200 IU/day vs. no offer (95% CI)** | **P** | **P for interaction for 3200 IU/day vs. no offer (95% CI)** | **Odds ratio for 800 IU/day vs. no offer (95% CI)** | **P** | **P for interaction for 800 IU/day vs. no offer (95% CI)** |
| --- | --- | --- | --- | --- | --- | --- | --- | --- | --- | --- |
| Proportion of all participants developing test-confirmed COVID-19^(1)^ (%) | Unvaccinated | 0/68  (0.0) | 5/69  (7.2) | 9/191  (4.7) | --^(2)^ | -- | --^3^ | 1.58 (0.51, 4.89) | 0.43 | 0.83 |
|  | Had ≥1 dose of COVID-19 vaccine | 32/1430  (2.2) | 43/1420  (3.0) | 59/2673  (2.2) | 1.01 (0.66, 1.57) | 0.95 |  | 1.38 (0.93, 2.06) | 0.11 |  |
| Proportion of participants developing test-confirmed COVID-19 who reported symptoms lasting more than four weeks^2^ (%) | Unvaccinated | 0/0 (0.0) | 2/5  (40.0) | 2/9  (22.2) | --^(2)^ | -- | --^3^ | 2.33 (0.22, 25.2) | 0.49 | 0.89 |
|  | Had ≥1 dose of COVID-19 vaccine | 11/32  (34.4) | 19/43  (44.2) | 17/59 (28.8) | 1.29 (0.51, 3.25) | 0.58 |  | 1.96 (0.86, 4.46) | 0.11 |  |

1, confirmed by RT-PCR and/or antigen testing for SARS-CoV-2. 2, OR incalculable due to zero events among unvaccinated participants in the 3200 IU/day offer arm. 3, P for interaction incalculable due to zero events among unvaccinated participants in the 3200 IU/day offer arm.

### Table S6. Adverse Events, by Allocation

|  |  | **3200 IU/day offer (n=1,515)^(1)^** | | **800 IU /day offer (n=1,515)^(1)^** | | **No offer (n=2,949)^(1)^** | |
| --- | --- | --- | --- | --- | --- | --- | --- |
|  |  | **No. of Events** | **No. of Participants with ≥1 Event** | **No. of Events** | **No. of Participants with ≥1 Event** | **No. of Events** | **No. of Participants with ≥1 Event** |
| Death^(2)^ |  | 2 | 2 | 1 | 1 | 4 | 4 |
| Non-fatal serious adverse event^(2)^ |  | 97 | 85 | 95 | 85 | 164 | 143 |
| Non-serious adverse event leading to discontinuation of study supplement^(3)^ |  | 25 | 25 | 23 | 23 |  |  |
| Other monitored safety conditions | Hypercalcemia^(4)^ | 4 | 4 | 0 | 0 | 0 | 0 |
|  | Hypervitaminosis D^(5)^ | 0 | 0 | 0 | 0 | 1 | 1 |
|  | Nephrolithiasis | 2 | 1 | 0 | 0 | 0 | 0 |

(1), denominator as per intention to treat analysis, i.e., participants with follow-up data from at least one timepoint. (2) details in Appendix Table S4. (3) details in Appendix Table S5. (4) defined as serum corrected calcium concentration >2.65 mmol/L. (5) defined as 25(OH)D concentration >220 nmol/L.

### Table S7: Line Listing of Serious Adverse Events, by Allocation

| **Type of SAE** | **Diagnosis** | **Events in 3200 IU/day arm, N**^(1)^ | **Events in 800 IU/day arm, N**^(2)^ | **Events in no offer arm, N**^(3)^ |
| --- | --- | --- | --- | --- |
| Death | Cervical cancer | 0 | 1 | 0 |
| Death | Deep vein phlebitis | 0 | 0 | 1 |
| Death | Lung cancer | 0 | 0 | 1 |
| Death | Myocardial infarction | 1 | 0 | 0 |
| Death | Pancreatic cancer | 1 | 0 | 0 |
| Death | Unknown cause | 0 | 0 | 2 |
| Hospitalisation | Abdominal pain, cause unknown | 0 | 1 | 0 |
| Hospitalisation | Acute coronary syndrome | 6 | 4 | 2 |
| Hospitalisation | Addisonian crisis | 0 | 0 | 1 |
| Hospitalisation | Allergic reaction | 0 | 1 | 0 |
| Hospitalisation | Anaemia | 1 | 1 | 0 |
| Hospitalisation | Anal fistula | 0 | 1 | 4 |
| Hospitalisation | Anxiety/depression | 0 | 1 | 0 |
| Hospitalisation | Appendicitis | 1 | 1 | 3 |
| Hospitalisation | Arthritis | 0 | 1 | 0 |
| Hospitalisation | Arthroscopy | 1 | 1 | 1 |
| Hospitalisation | Ascending aortic replacement | 0 | 1 | 1 |
| Hospitalisation | Bladder cancer | 0 | 1 | 1 |
| Hospitalisation | Bone cyst removal | 2 | 0 | 1 |
| Hospitalisation | Bone graft | 1 | 0 | 0 |
| Hospitalisation | Bowel obstruction | 0 | 1 | 1 |
| Hospitalisation | Brain tumour | 1 | 0 | 0 |
| Hospitalisation | Breast cancer | 0 | 0 | 1 |
| Hospitalisation | Bunionectomy | 0 | 1 | 2 |
| Hospitalisation | Cancer, unknown primary | 1 | 0 | 0 |
| Hospitalisation | Cardiac arrhythmia | 3 | 1 | 6 |
| Hospitalisation | Cardiac failure | 0 | 1 | 0 |
| Hospitalisation | Cellulitis | 0 | 0 | 1 |
| Hospitalisation | Cerebrovascular accident | 2 | 4 | 4 |
| Hospitalisation | Chest pain, cause unknown | 1 | 0 | 0 |
| Hospitalisation | Cholecystectomy | 1 | 1 | 2 |
| Hospitalisation | Cholecystitis | 0 | 1 | 0 |
| Hospitalisation | Churg-Strauss syndrome | 1 | 0 | 2 |
| Hospitalisation | Colon cancer | 2 | 0 | 3 |
| Hospitalisation | Colonoscopy | 1 | 2 | 4 |
| Hospitalisation | Constipation | 1 | 0 | 2 |
| Hospitalisation | Cough, cause unknown | 0 | 1 | 3 |
| Hospitalisation | COVID-19 | 29 | 24 | 40 |
| Hospitalisation | Crohn's disease | 0 | 0 | 1 |
| Hospitalisation | Dehydration | 1 | 0 | 0 |
| Hospitalisation | Dental procedure | 0 | 0 | 2 |
| Hospitalisation | Diabetic ketoacidosis | 0 | 0 | 1 |
| Hospitalisation | Duodenal resection | 0 | 1 | 0 |
| Hospitalisation | Dyspnoea, cause unknown | 1 | 0 | 1 |
| Hospitalisation | Endometriosis | 0 | 0 | 1 |
| Hospitalisation | Excision basal cell carcinoma | 1 | 2 | 0 |
| Hospitalisation | Excision breast lump | 0 | 1 | 0 |
| Hospitalisation | Fall | 0 | 2 | 1 |
| Hospitalisation | Femoroacetabular impingement | 0 | 0 | 1 |
| Hospitalisation | Fracture | 1 | 6 | 6 |
| Hospitalisation | Fracture plate removal | 1 | 0 | 0 |
| Hospitalisation | Functional neurological disorder | 1 | 0 | 2 |
| Hospitalisation | Gastrointestinal haemorrhage | 1 | 0 | 1 |
| Hospitalisation | Gastro-oesophageal reflux disease | 0 | 1 | 1 |
| Hospitalisation | Haematuria | 0 | 0 | 1 |
| Hospitalisation | Head injury | 0 | 3 | 2 |
| Hospitalisation | Hernia repair | 0 | 0 | 1 |
| Hospitalisation | Hypertension | 0 | 1 | 1 |
| Hospitalisation | Hypoglycaemia | 1 | 0 | 0 |
| Hospitalisation | Hysteroscopy | 0 | 0 | 1 |
| Hospitalisation | Intestinal obstruction | 1 | 0 | 0 |
| Hospitalisation | Joint replacement | 2 | 1 | 12 |
| Hospitalisation | Lens replacement surgery | 3 | 1 | 2 |
| Hospitalisation | Lymphoma | 0 | 1 | 0 |
| Hospitalisation | Manipulation under anaesthesia | 0 | 0 | 2 |
| Hospitalisation | Migraine | 0 | 1 | 1 |
| Hospitalisation | Miscarriage | 0 | 2 | 1 |
| Hospitalisation | Nephrolithiasis | 2 | 0 | 0 |
| Hospitalisation | Non-COVID-19 acute respiratory infection | 11 | 7 | 12 |
| Hospitalisation | Ovarian cystectomy | 0 | 0 | 2 |
| Hospitalisation | Pancreatic cancer | 2 | 0 | 0 |
| Hospitalisation | Pancreatitis | 1 | 0 | 0 |
| Hospitalisation | Percutaneous coronary angiography | 0 | 0 | 1 |
| Hospitalisation | Perianal fistula | 0 | 0 | 1 |
| Hospitalisation | Peritonitis | 0 | 0 | 1 |
| Hospitalisation | Pleurisy | 0 | 1 | 0 |
| Hospitalisation | Pneumothorax | 0 | 0 | 2 |
| Hospitalisation | Prostatic hypertrophy | 0 | 1 | 0 |
| Hospitalisation | Pulmonary thrombo-embolism | 0 | 0 | 1 |
| Hospitalisation | Renal failure | 1 | 0 | 0 |
| Hospitalisation | Road traffic accident | 0 | 0 | 1 |
| Hospitalisation | Sciatica | 0 | 1 | 0 |
| Hospitalisation | Scrotal hydrocele | 0 | 1 | 0 |
| Hospitalisation | Seizure | 0 | 0 | 2 |
| Hospitalisation | Sepsis | 2 | 0 | 1 |
| Hospitalisation | Shingles | 0 | 0 | 1 |
| Hospitalisation | Shoulder pain, cause unknown | 1 | 0 | 0 |
| Hospitalisation | Skin biopsy | 0 | 1 | 0 |
| Hospitalisation | Soft tissue injury | 2 | 0 | 4 |
| Hospitalisation | Spinal disc replacement | 1 | 0 | 0 |
| Hospitalisation | Spinal nerve entrapment | 0 | 2 | 0 |
| Hospitalisation | Sudden sensorineural hearing loss | 0 | 1 | 0 |
| Hospitalisation | Syncope | 0 | 2 | 1 |
| Hospitalisation | Testicular torsion | 0 | 1 | 0 |
| Hospitalisation | Thyroidectomy | 0 | 0 | 1 |
| Hospitalisation | Torticollis | 1 | 0 | 0 |
| Hospitalisation | Urinary retention | 0 | 1 | 1 |
| Hospitalisation | Urinary tract infection | 0 | 1 | 2 |
| Hospitalisation | Uterine polyp | 1 | 0 | 1 |
| Hospitalisation | Uterine prolapse | 1 | 0 | 0 |
| Hospitalisation | Vaginal haemorrhage | 1 | 1 | 2 |
| Hospitalisation | Vomiting | 1 | 0 | 0 |
| Total |  | 99 | 96 | 168 |

1, these events arose in 87 participants randomised to the 3200 IU/day arm

2, these events arose in 86 participants randomised to the 800 IU/day arm

3, these events arose in 147 participants randomised to the no offer arm

### Table S8: Non-serious adverse events leading to discontinuation of study supplements in either intervention arm, by allocation.

| **Nature of adverse event** | **Number of events arising in 3200 IU/day arm** | **Number of events arising in 800 IU/day arm** |
| --- | --- | --- |
| Hypercalcemia | 4 | 0 |
| Gastro-intestinal symptoms* | 16 | 16 |
| General malaise | 1 | 4 |
| Headache | 1 | 2 |
| Rash | 2 | 1 |

^*e.g., nausea, dyspepsia, constipation^

### Figure S1: Imputed vs. measured blood 25-hydroxyvitamin D concentrations at baseline.

Values were imputed for participants randomised to no offer and measured for participants randomised to either intervention arm. Bars show mean and standard deviation for each group.

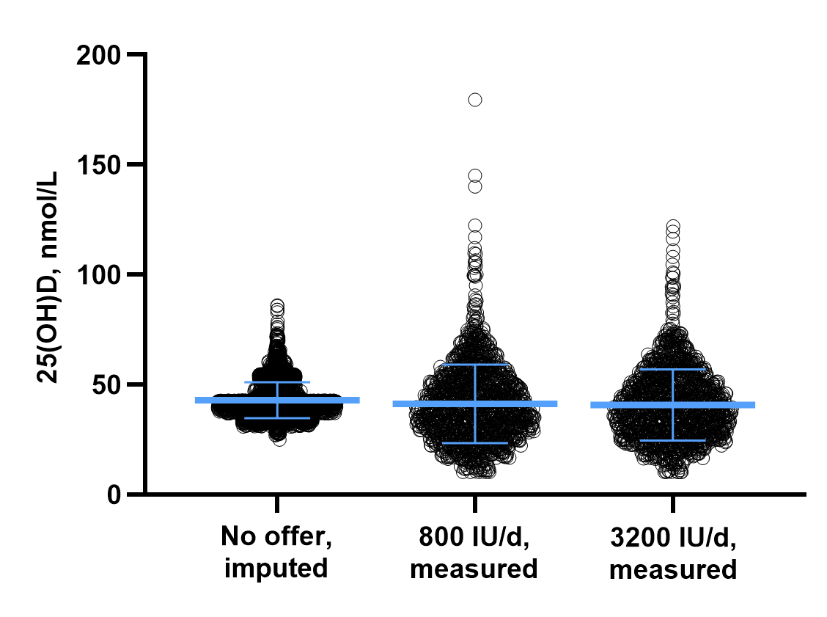

### Figure S2: Cumulative hazard plots showing time to COVID-19 by allocation, stratified by SARS-CoV-2 vaccination status.

**A**, vaccine-naïve participants randomised to lower-dose vs. no offer. **B**, vaccinated participants randomised to lower-dose vs. no offer. **C**, vaccine-naïve participants randomised to higher-dose vs. no offer. **D**, vaccinated participants randomised to higher-dose vs. no offer. Shaded areas show 95% confidence intervals (CI). HR, hazard ratio.

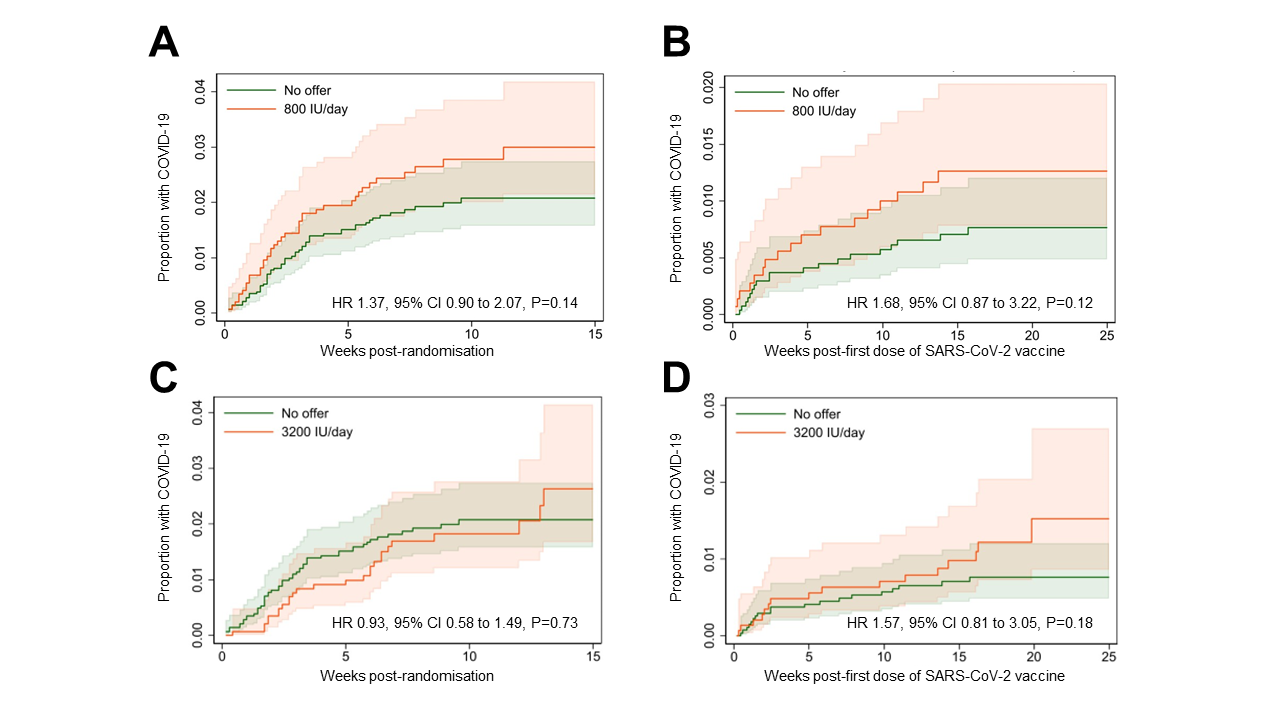
